# Analysis of presence and short-term persistence of SARS-CoV-2 neutralizing antibodies in COVID-19 convalescent plasma donors

**DOI:** 10.1101/2020.09.01.20185942

**Authors:** Kyle Annen, Thomas E. Morrison, Melkon G. DomBourian, Mary K. McCarthy, Leah Huey, Patricia Merkel, Gillian Andersen, Eileen Schwartz, Vijaya Knight

**Author notes:** Co-first authors.

## Abstract

In March 2020, the FDA approved the use of COVID-19 convalescent plasma (CCP) as an investigational new drug for treatment of COVID-19. Since then, collection of CCP from COVID-19 recovered patients has been implemented in several donor centers across the country. Children’s Hospital Colorado rapidly put into practice a CCP collection protocol, necessitating the development and implementation of assays to evaluate SARS-CoV-2 antibodies in CCP units. We evaluated 87 separate units of CCP collected from 36 donors over two to four sequential donations using both antigen-binding assays for SARS-CoV-2 nucleoprotein and spike antigens, and a live virus focus reduction neutralization test (FRNT_50_). Our data shows that the majority of donors (83%) had a FRNT_50_ titer of 1:80 or greater, and 61% had a titer ≥ 1:160, which meet the FDA’s criteria for acceptable CCP units. Additionally, our data indicates that analysis of antibodies to a single SARS-CoV-2 antigen is likely to miss a percentage of seroconverters; however, these individuals tend to have neutralizing antibody titers of <1:80. Of note, there was considerable variability in the short term, sustained antibody response, measured by neutralizing antibody titers, among our donor population.

## Introduction

The Food and Drug Administration (FDA) authorized the use of COVID-19 convalescent plasma (CCP) as an investigational new drug for the treatment of COVID-19 (1).

Initially, convalescent plasma donors were accepted only with a confirmed SARS-CoV-2 positive PCR. Donors were required to be symptom-free for at least 14 days prior to donation, SARS-CoV-2 PCR negative upon subsequent testing, and meet all other blood donation eligibility requirements (2). The most recent guidance dropped the requirement for a negative diagnostic test to qualify donors and describes an additional pathway for CCP donors to be deemed eligible to donate: a confirmed positive SARS- CoV-2-specific IgG based serology test (1).

During the early stages of the COVID-19 crises in the U.S., Children’s Hospital of Colorado (CHCO) rapidly developed and implemented protocols for the collection of CCP. Since our first collection on March 31, 2020, we have collected 548 CCP units. During the beginning phase of CCP collection, a primary challenge was identifying donors who met the SARS-CoV-2 PCR positive test and symptom resolution criteria (i.e., asymptomatic for 14 days or more). Although the FDA previously established that plasma for the purpose of transfusion be collected no more frequently than every 28 days (3), the FDA has allowed for more frequent blood collection at the discretion of the donation center medical director (personal correspondence Peter Marks, April 16, 2020). This exception could significantly improve access to CCP by allowing for an earlier return of donors who had previously been successfully screened and tested. Since coagulation factors, such as fibrinogen and albumin, are expected to be replaced in a donor’s plasma within one week(4), several donation centers in the country, including CHCO, allowed repeat convalescent plasma donors as frequently as every 7 days. However, the impact on the donors anti-SARS-CoV-2 antibody levels with this frequency of donation, or any frequency, is unknown, as is the pattern of decline or retention of antibodies to SARS-CoV-2.

A number of assays for detection of SARS-CoV-2 antibodies, including ELISA, high throughput immunoassay platforms, and rapid detection lateral flow assays became available in late March, enabling relatively rapid screening of CCP units for the presence of anti-SARS-CoV-2 antibodies, commonly to either the SARS-CoV-2 nucleocapsid or spike antigens, such as the S1 and receptor binding domain (RBD) of the spike protein (5, 6). However, FDA recommendations state that CCP units that are eligible for transfusion into COVID-19 patients should have a neutralizing antibody titer of at least 1:80, and preferably 1:160 (2). This guidance poses challenges for laboratories that are screening CCP units for SARS-CoV-2 antibodies with antigen binding assays, many of which are primarily qualitative in nature, and do not provide information regarding potential for SARS-CoV-2 neutralization. Therefore, a comparison of antigen binding assays with virus neutralizing antibody titer is increasingly important to enable triage of CCP units and to develop criteria for units that are suitable for transfusion into patients with COVID-19.

Here, we compare two ELISA assays, both currently implemented in clinical laboratories for clinical diagnostics and for screening of CCP, with a SARS-CoV-2 virus neutralization assay in our CCP donor population. As CCP donors are currently permitted to return for relatively frequent plasma donations, we have additionally examined the persistence of SARS-CoV-2 antibodies using the neutralizing antibody assay. These data contribute to our understanding of the neutralizing antibody response to SARS-CoV-2, its correlation with development of N and S1 binding antibodies, persistence of the neutralizing antibody response, and ultimately strengthen the criteria for CCP donors and analysis of CCP as a therapeutic for COVID-19.

## Materials and Methods

### COVID-19 convalescent plasma donors

SARS-CoV-2 PCR positive individuals who were eligible to donate plasma according to FDA criteria for CCP donors were enrolled under the CHCO CCP donor program. Aliquots of plasma and serum were stored at -80°C until analysis.

### SARS-CoV-2 IgG ELISA

Two commercial ELISAs, CE-marked Epitope Diagnostics Inc. (EDI, San Diego, CA) and Euroimmun (both CE-marked and FDA EUA approved, Lubeck, Germany) were utilized in this study. Both of the ELISAs report results qualitatively, based on a single dilution. The EDI ELISA utilizes a SARS-CoV-2 recombinant nucleocapsid (N) protein as the coating antigen. Positive and negative assay controls and samples were diluted 1:100 with the kit-specific COVID-19 IgG sample diluent and added to the wells, followed by a 30 min incubation at room temperature. Plates were washed five times using the kit-specific wash buffer and anti-human IgG horse radish peroxidase (HRP)-conjugated detection antibody was added, followed by a 30 min incubation. Plates were washed five times, and the signal was developed using tetramethylbenzidine (TMB). Absorbance was read at 450 nm within 10 min of halting the reaction.

The Euroimmun ELISA assay utilizes the S1 domain, which includes the receptor binding domain (RBD) of the SARS-CoV-2 spike protein (7). For this assay, a kit-specific calibrator, positive and negative controls and samples, were diluted 1:101 with the kit-specific dilution buffer and added to pre-coated wells. Following a 1 h incubation at 37°C, plates were washed three times with kit-specific wash buffer. Anti-human IgG-HRP conjugated detection antibody was added and plates were incubated for 30 min at 37°C followed by three washes. TMB was added and absorbance read at 450 nm within 10 min of halting the reaction.

### Interpretation of ELISA results

EDI ELISA: Positive, negative and borderline results were calculated based on the average optical density (OD) value for the negative control assayed in triplicate for the specific assay. The positive and negative cut-off values were calculated using the formula: positive cut-off = 1.1 x (xNC+0.18) and negative cut-off = 0.9 x (xNC + 0.18), where xNC is the average OD_450_ of triplicate negative control OD_450_ values. Samples that had OD values between positive and negative cut-off values were reported as borderline.

Euroimmun ELISA: The ratio of the sample OD_450_ values to the calibrator OD_450_ values was calculated for all samples and controls. Samples with a ratio of less than 0.8 were reported as negative, samples with a ratio of greater than 1.1 were reported as positive, and ratios between 0.8 and 1.1 were reported as borderline.

### Focus Reduction Neutralization Assay (FRNT)

Vero E6 cells (ATCC, Manassas, VA) were seeded in 96-well plates. Serum samples were heat inactivated and serially diluted (2-fold, starting at 1:10) in DMEM (ThermoFisher, Pittsburgh, PA, USA) plus 1% FBS (manufacturer, city, state) in 96-well plates. Approximately 100 focus-forming units (FFU) of SARS-CoV-2 USA-WA1/2020 (8)(deposited by the Centers for Disease Control and Prevention and obtained through BEI Resources, NIAID, NIH) was added to each well and the serum plus virus mixture was incubated for 1 h at 37°C. At the end of 1 h, medium was removed from cells and the serum sample plus virus mixture was added for 1 h at 37°C. After 1 h, samples were removed and cells overlaid with 1% methylcellulose (MilliporeSigma, St. Louis, MO) in MEM (ThermoFisher, Pittsburgh, PA, USA)/2% FBS and incubated 30 h at 37°C. Cells were fixed with 4% paraformaldehyde (Acros Organics, Pittsburgh, PA, USA) and probed with 1 μg/mL of an anti-SARS-CoV spike monoclonal antibody (CR3022, Absolute Antibody, Boston, MA, USA) in Perm Wash (1X PBS/0.1% saponin/0.1% bovine serum albumin [BSA]) for 2 h at RT. After washing, cells were incubated with horseradish peroxidase (HRP)-conjugated goat anti-human IgG (Southern Biotech, Birmingham, AL, USA, 1:1,000) for 1.5 h at RT. After washing, SARS-CoV-2-positive foci were visualized with TrueBlue substrate (ThermoFisher, Pittsburgh, PA, USA) and counted using a CTL Biospot analyzer and Biospot software (Cellular Technology Ltd, Shaker Heights, OH, USA). The FRNT_50_ titer was calculated relative to a virus only control (no serum) set at 100%, using GraphPad Prism 8 (La Jolla, CA, USA) default nonlinear curve fit constrained between 0 and 100%.

### Data analysis

Data were analyzed using GraphPad Prism Version 8 (San Diego, CA) and Microsoft Excel 2016 (Microsoft, Redmond, WA). For OD_450_ values and S1-RBD ratios, the mean and 95% confidence intervals were calculated using GraphPad Prism’s statistical analysis package. Significant differences between groups were calculated using Welch’s test for unequal variances. The difference between groups was considered statistically significant when p<0.05.

## Results

### Characterization of donor serum samples

Eighty-seven samples from thirty-six CCP donors from the CHCO Blood Donor Center were included in this study. All plasma donors made an initial donation 12-42 days following a positive SARS-CoV-2 PCR result, and again at the intervals shown in Table 1. Of the thirty-six donors, twenty-four donated plasma twice, nine donors, three times, and three donors, four times. The intervals between sequential plasma donations ranged from 7 to 24 days.

**Table 1.** Interval (days) between CCP collection for individual donors.

| Donor ID | Interval between positive PCR result and initial donation | Interval between initial and second donation | Interval between second and third donation | Interval between third and fourth donation |
| --- | --- | --- | --- | --- |
| 001-D | 33 | 7 |  |  |
| 002-D | 36 | 7 |  |  |
| 003-D | 28 | 8 | 17 |  |
| 005-D | 29 | 7 | 9 |  |
| 006-D | 29 | 7 | 9 |  |
| 007-D | 42 | 8 | 6 | 8 |
| 008-D | 20 | 19 |  |  |
| 009-D | 18 | 12 | 8 | 7 |
| 010-D | 33 | 18 |  |  |
| 011-D | 30 | 7 |  |  |
| 012-D | 19 | 13 | 7 |  |
| 013-D | 31 | 8 | 14 |  |
| 014-D | 25 | 13 | 11 |  |
| 015-D | 22 | 11 | 7 | 7 |
| 016-D | 36 | 8 | 9 |  |
| 018-D | 27 | 15 |  |  |
| 019-D | 26 | 8 |  |  |
| 020-D | 22 | 11 | 7 |  |
| 021-D | 32 | 8 |  |  |
| 022-D | 24 | 11 |  |  |
| 023-D | 31 | 8 |  |  |
| 024-D | 33 | 12 |  |  |
| 025-D | 29 | 12 |  |  |
| 026-D | 27 | 8 |  |  |
| 027-D | 26 | 20 |  |  |
| 028-D | 12 | 14 |  |  |
| 029-D | 36 | 5 |  |  |
| 030-D | 23 | 18 |  |  |
| 031-D | 28 | 11 |  |  |
| 032-D | 24 | 14 |  |  |
| 033-D | 22 | 24 |  |  |
| 034-D | 29 | 12 |  |  |
| 035-D | 36 | 9 |  |  |
| 036-D | 24 | 10 |  |  |
| 037-D | 26 | 9 | 13 |  |

### Comparison of N and S1-RBD antibody detection with virus neutralizing activity

To determine if qualitative IgG antibody detection by ELISA, whether against the N or the S1-RBD antigen, correlated with virus neutralizing activity, samples were analyzed for the presence of anti-N IgG antibodies using the Epitope Diagnostics ELISA, anti-S1- RBD IgG antibodies using the Euroimmun ELISA, and neutralizing activity using a live virus focus reduction assay. Samples with a neutralizing antibody titer of 1:80 or greater had a positive or borderline-positive result for both N and S1-RBD IgG antibodies, with the exception of sample 018-D (neutralizing titer of 1:85), which was positive for N and negative for S1 -RBD, and sample 023-D (neutralizing titer of 1:91) which was negative for N and was not analyzed for S1-RBD antibodies (Table 2). More variability between anti-N and anti-S1-RBD IgG ELISA results was noted for samples with lower neutralizing antibody titers, particularly for the five samples that had neutralizing titers of <1:40. These samples were negative for anti-N IgG antibodies; four of the five samples were either negative or borderline-positive for anti-S1-RBD IgG, and one was positive. Of note, donor 019-D (samples 1 and 2) remained persistently negative for anti-N IgG, had a marginal increase in anti-S1-RBD IgG on the second CCP donation, and had very low FRNT50 titers.

**Table 2.**
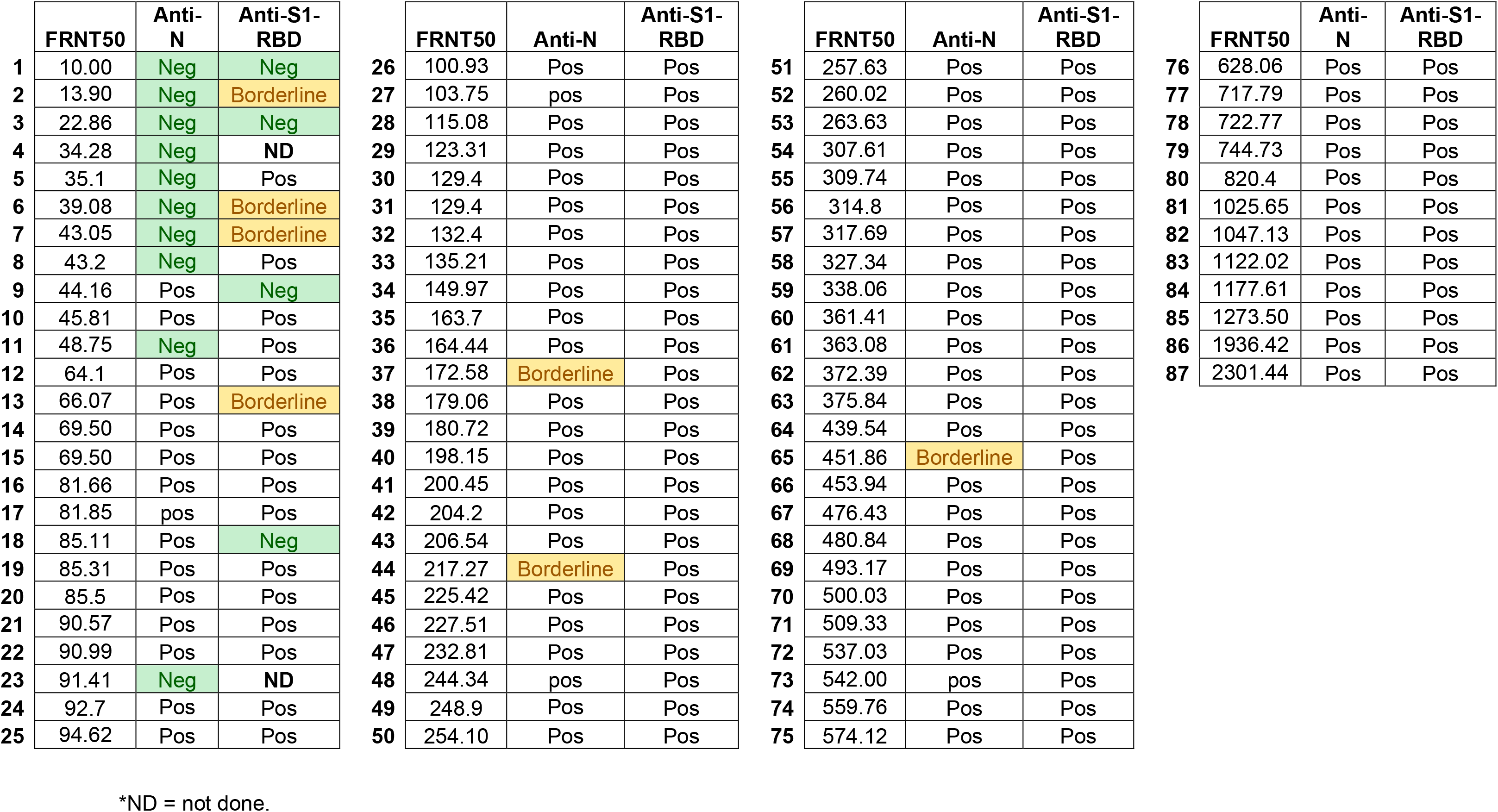
Correlation of FRNT_50_ reciprocal titers and anti-N IgG or anti-S1-RBD IgG qualitative results.

|  | FRNT50 | Anti-N | Anti-S1-RBD |  | FRNT50 | Anti-N | Anti-S1-RBD |  | FRNT50 | Anti-N | Anti-S1-RBD |  | FRNT50 | Anti-N | Anti-S1-RBD |
| --- | --- | --- | --- | --- | --- | --- | --- | --- | --- | --- | --- | --- | --- | --- | --- |
| 1 | 10.00 | Neg | Neg | 26 | 100.93 | Pos | Pos | 51 | 257.63 | Pos | Pos | 76 | 628.06 | Pos | Pos |
| 2 | 13.90 | Neg | Borderline | 27 | 103.75 | pos | Pos | 52 | 260.02 | Pos | Pos | 77 | 717.79 | Pos | Pos |
| 3 | 22.86 | Neg | Neg | 28 | 115.08 | Pos | Pos | 53 | 263.63 | Pos | Pos | 78 | 722.77 | Pos | Pos |
| 4 | 34.28 | Neg | ND | 29 | 123.31 | Pos | Pos | 54 | 307.61 | Pos | Pos | 79 | 744.73 | Pos | Pos |
| 5 | 35.1 | Neg | Pos | 30 | 129.4 | Pos | Pos | 55 | 309.74 | Pos | Pos | 80 | 820.4 | Pos | Pos |
| 6 | 39.08 | Neg | Borderline | 31 | 129.4 | Pos | Pos | 56 | 314.8 | Pos | Pos | 81 | 1025.65 | Pos | Pos |
| 7 | 43.05 | Neg | Borderline | 32 | 132.4 | Pos | Pos | 57 | 317.69 | Pos | Pos | 82 | 1047.13 | Pos | Pos |
| 8 | 43.2 | Neg | Pos | 33 | 135.21 | Pos | Pos | 58 | 327.34 | Pos | Pos | 83 | 1122.02 | Pos | Pos |
| 9 | 44.16 | Pos | Neg | 34 | 149.97 | Pos | Pos | 59 | 338.06 | Pos | Pos | 84 | 1177.61 | Pos | Pos |
| 10 | 45.81 | Pos | Pos | 35 | 163.7 | Pos | Pos | 60 | 361.41 | Pos | Pos | 85 | 1273.50 | Pos | Pos |
| 11 | 48.75 | Neg | Pos | 36 | 164.44 | Pos | Pos | 61 | 363.08 | Pos | Pos | 86 | 1936.42 | Pos | Pos |
| 12 | 64.1 | Pos | Pos | 37 | 172.58 | Borderline | Pos | 62 | 372.39 | Pos | Pos | 87 | 2301.44 | Pos | Pos |
| 13 | 66.07 | Pos | Borderline | 38 | 179.06 | Pos | Pos | 63 | 375.84 | Pos | Pos |  |  |  |  |
| 14 | 69.50 | Pos | Pos | 39 | 180.72 | Pos | Pos | 64 | 439.54 | Pos | Pos |  |  |  |  |
| 15 | 69.50 | Pos | Pos | 40 | 198.15 | Pos | Pos | 65 | 451.86 | Borderline | Pos |  |  |  |  |
| 16 | 81.66 | Pos | Pos | 41 | 200.45 | Pos | Pos | 66 | 453.94 | Pos | Pos |  |  |  |  |
| 17 | 81.85 | pos | Pos | 42 | 204.2 | Pos | Pos | 67 | 476.43 | Pos | Pos |  |  |  |  |
| 18 | 85.11 | Pos | Neg | 43 | 206.54 | Pos | Pos | 68 | 480.84 | Pos | Pos |  |  |  |  |
| 19 | 85.31 | Pos | Pos | 44 | 217.27 | Borderline | Pos | 69 | 493.17 | Pos | Pos |  |  |  |  |
| 20 | 85.5 | Pos | Pos | 45 | 225.42 | Pos | Pos | 70 | 500.03 | Pos | Pos |  |  |  |  |
| 21 | 90.57 | Pos | Pos | 46 | 227.51 | Pos | Pos | 71 | 509.33 | Pos | Pos |  |  |  |  |
| 22 | 90.99 | Pos | Pos | 47 | 232.81 | Pos | Pos | 72 | 537.03 | Pos | Pos |  |  |  |  |
| 23 | 91.41 | Neg | ND | 48 | 244.34 | pos | Pos | 73 | 542.00 | pos | Pos |  |  |  |  |
| 24 | 92.7 | Pos | Pos | 49 | 248.9 | Pos | Pos | 74 | 559.76 | Pos | Pos |  |  |  |  |
| 25 | 94.62 | Pos | Pos | 50 | 254.10 | Pos | Pos | 75 | 574.12 | Pos | Pos |  |  |  |  |

\*ND = not done.

To determine whether OD_450_ values for anti-N IgG antibodies or the ratio for anti-S1- RBD IgG antibodies was predictive of neutralizing antibody titers, we compared the numerical values associated with a positive or negative ELISA result with the corresponding neutralizing antibody titer (Figure 1). Samples with a neutralizing antibody titer of <1:80 had the lowest OD_450_ values for N or ratios for S1-RBD. An increase in OD_450_ value or an increase in S1-RBD ratio independently correlated with a significant increase in neutralizing antibody titers (Figures 1A and 1B). However, given the wide variability among OD_450_ values and ratios within each group, we performed a 3way comparison of the data to examine whether there was a relationship between the level of positivity for anti-N IgG, anti-S1-RBD IgG, and neutralizing antibody titers (Figure 2). Two of the samples with the highest neutralizing antibody titers, both from donor 001 -D, had the highest ratios for anti-S1-RBD IgG, and although they were strongly positive for anti-N IgG antibodies, they did not have the highest OD_450_ values in this sample set. In general, samples with both low anti-N IgG and anti-S1-RBD IgG numerical values correlated well with low or minimal neutralizing activity. Five samples with neutralizing titers between 1:1,000-1:1,500 had varying levels of positivity for anti-N IgG and anti-S1-RBD IgG, and three samples with the highest OD_450_ values for anti-N IgG antibodies had neutralizing antibody titers in the range of 1:200 (Figure 2).

**Figure 1.**
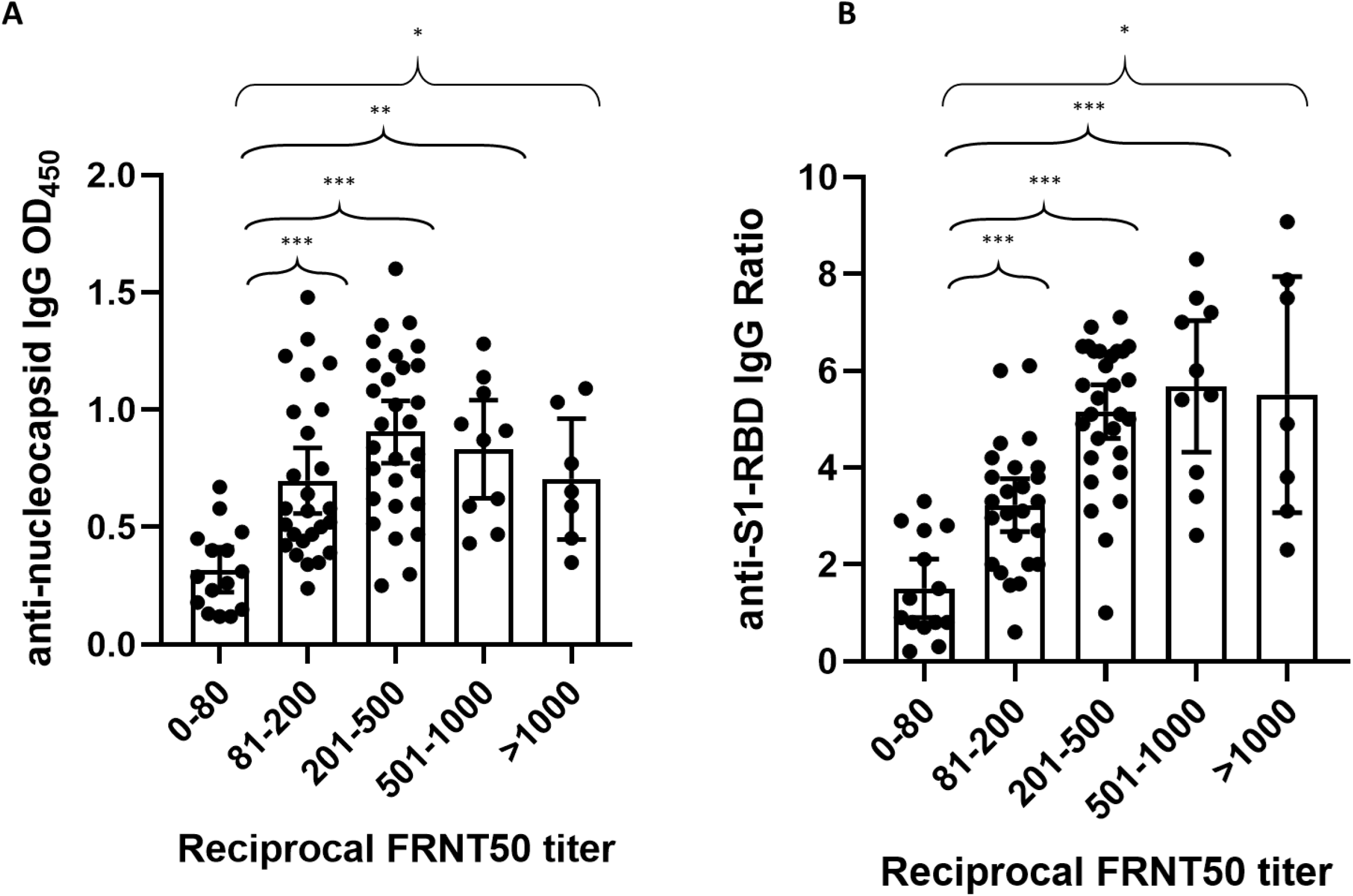
Comparison of reciprocal FRNT50 titer with either anti-nucleocapsid IgG (A) or anti-SI-RBD IgG (B). Data are the mean and 95% confidence intervals for each group of FRNT50 titers. N=86 for FRNT50 vs anti-N antibodies and N=84 for FRNT50 vs anti- S1-RBD antibodies. *=p<0.05, **=p<0.001, and ***=p<0.0001 using Welch’s test for unequal variance.

**Figure 2:**
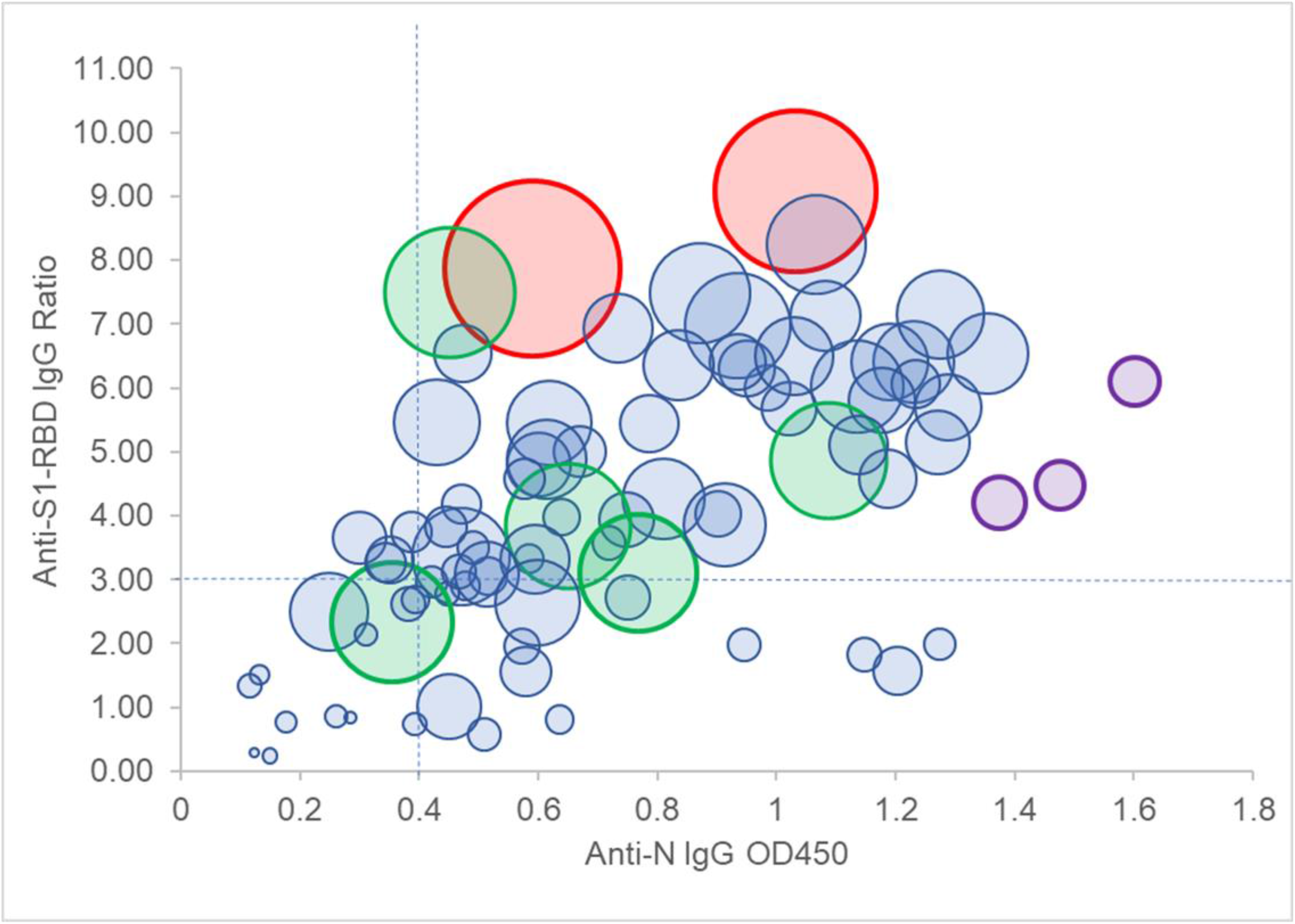
Comparison of anti-nucleocapsid IgG (OD_450_, x-axis), anti-S1-RBD IgG (ratio, y-axis), and FRNT50 reciprocal titer (relative size of bubble, larger bubbles correspond to higher titers). Eighty-five samples were compared for correlation among anti-N, anti- S1-RBD and neutralizing antibody titers. Two samples with the highest FRNT50 titers (red circles), also had the highest S1-RBD ratios and moderately high levels of anti-N1. Three samples with the highest anti-N OD_450_ values had FRNT50 values of approximately 1:200 (purple circles), while five samples (green circles) had FRNT50 values of approximately 1:1000 but had varying levels of anti-N and anti-S1-RBD antibodies. Dashed lines indicate cut-off values for OD_450_ and ratios above which 90% of FRNT50 values were 80 or greater.

Given that these ELISAs were performed using a single dilution of serum, it is possible that the numerical values obtained are not in the linear range, and, therefore, these results affect correlations with neutralizing antibody titers as the FRNT50 is a quantitative assay. Nevertheless, an anti-N IgG OD_450_ of 0.4 and above correlated well with a neutralizing titer of ≥1:80 in 90% of the samples, and an anti-S1-RBD IgG ratio of 3.0 and above correlated with ≥1:80 neutralizing antibody titer in 82% of the samples (Table 3 and Figure 2). Only three samples with a neutralizing titer of ≥1:80 had an anti-N IgG OD_450_ <0.4 and an anti-S1-RBD IgG ratio <3.0. In general, when the anti-N IgG OD_450_ value was <0.4, the anti-S1-RBD IgG ratio was >3.0 and vice versa, indicating that a combination of the two assays accurately captured 96% of CCP samples with ≥1:80 neutralizing activity. Additionally, specificity of the anti-S1-RBD IgG ratio was greater for neutralizing titers, as 93% of samples with <1:80 neutralizing activity had <3.0 anti-S1-RBD IgG ratios, whereas greater than a quarter (27%) of these samples had anti-N IgG OD_450_ values of >0.4 (Table 3).

**Table 3:**
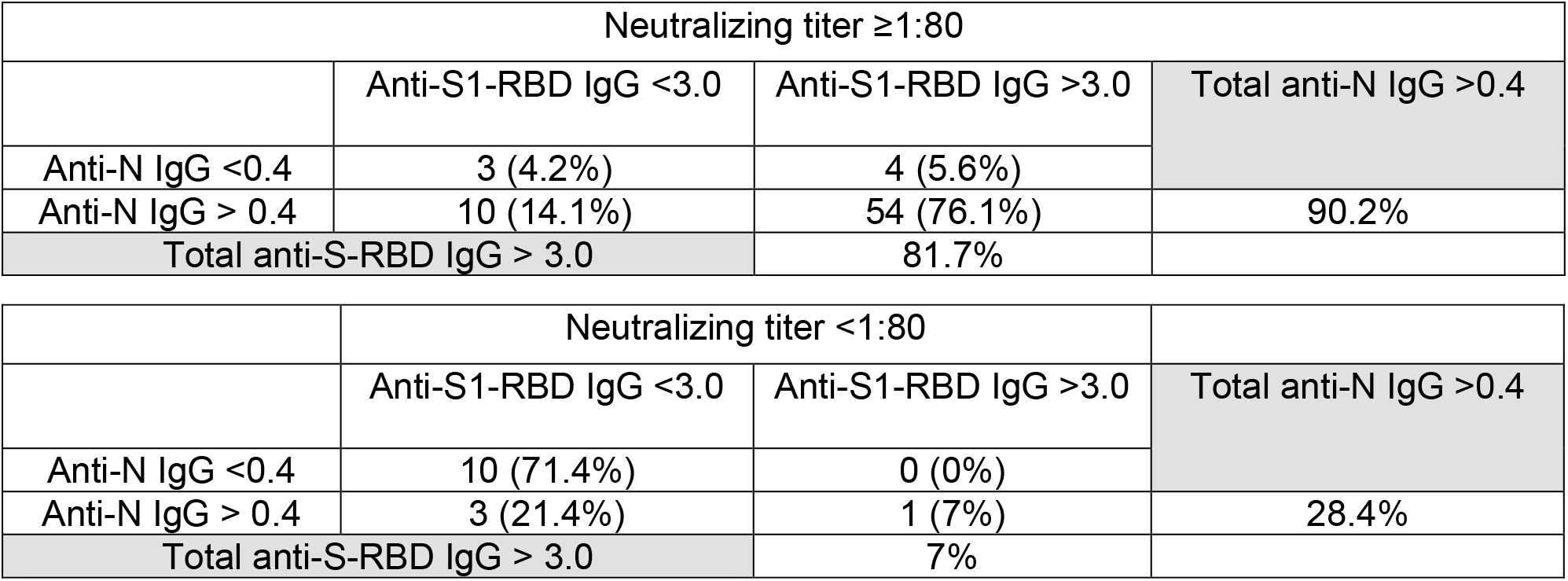
Correlation of >1:80 or <1:80 neutralizing antibody titers with anti-N or anti-S1-RBD level of positivity.

As recommended by the FDA, CCP units eligible for therapeutic use for COVID-19 patients are expected to have a neutralizing antibody titer of at least 1:80 and preferably ≥1:160. Of the 87 samples tested in this study, 72/87 (82.7%) had a neutralizing titer of ≥1:80 and 53/87 (60.9%) had a titer of ≥1:160 (Table 2 and Figure 3A).

**Figure 3.**
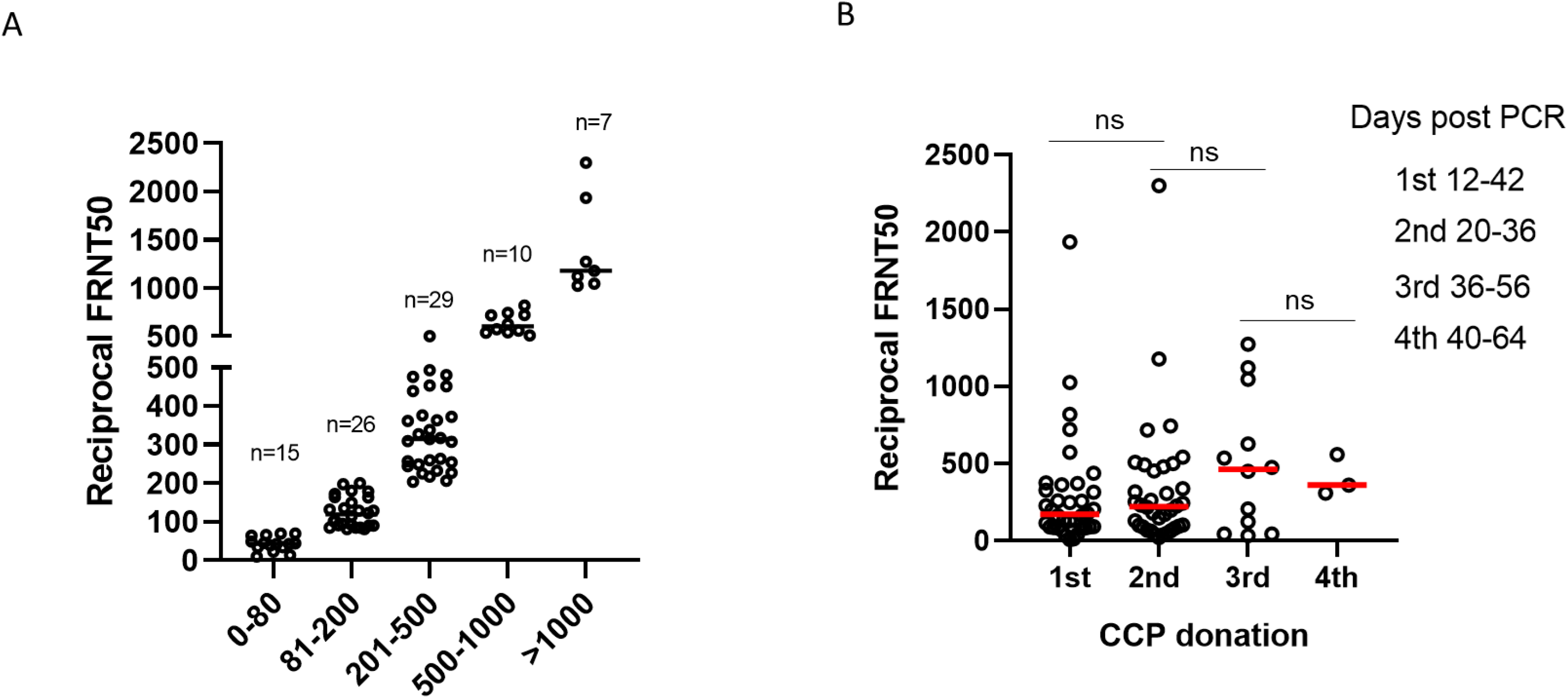
FRNT50 titers in CCP donors (A) and FRNT50 titers grouped by sequential CCP donations (B). Eighty-seven samples were analyzed for FRNT50 titers. Data are grouped by FRNT50 titer and the number of samples in each group is indicated (A). FRNT50 titers correlated with CCP collection time point. The difference between groups was considered significant (p<0.05) using Welch’s test for unequal variances. ns=not significant.

Neutralizing antibody titers were, in general, between 1/80 and 1/500 for the majority of samples tested (52%). Approximately 20% had titers greater than 1/500 and very few (7 of the 87 tested) had neutralizing antibody titers of >1:1000 (Figure 3A).

### Sustainability of the antibody response

Because analysis of N and S1-RBD IgG antibodies by single dilution ELISA is qualitative at best, we chose to analyze the robustness and sustainability of the SARS-CoV-2 antibody response by analyzing neutralizing antibody titers in sequential samples from the 36 donors included in this study. Neutralizing antibody titers at the time of initial donation varied significantly from <1:10 to almost 1:2000 (Figure 3B). Aggregate analysis of plasma samples at the time of initial donation, or between seven and 24 days following initial collection, showed an increase from baseline at the time of the second donation and an average greater increase at the third donation; however, these increases were not statistically significant (Figure 3B). Although the mean neutralizing antibody titer appeared to decrease at the time of the fourth donation, there were too few samples to ensure statistical significance at this time point. Given that the considerable variability of SARS-CoV-2 neutralizing antibody titers between donors may also be confounded by the interval between donations and the initial neutralizing antibody titer, we analyzed longevity of neutralizing antibody responses for individual donors. The majority of individuals donated two plasma units 7-24 days apart (Figures 4A and B). Only one donor (001-D) had a baseline titer of >1/1500, which rose to >1/2000 at the time of the second donation (Figure 4A). Of the 24 donors who donated plasma twice, neutralizing antibody titers decreased in seven (Figure 4B), increased slightly (< 2-fold) or remained relatively unchanged in 12, and increased between 2-6-fold in five donors (Figure 4C). Of the 12 donors with three or four sequential donations (Figures 5A and B), antibody titers increased or remained relatively unchanged from the initial plasma donation for seven donors and declined in the remaining five. Donor 015-D had the greatest decline in titers, from a 5-fold increase at the second and third donation to a 2-fold increase over baseline at the fourth donation. In general, for the seven donors who had an increase in neutralizing antibody titer over time, the fold increase in titer was moderate, with the exception of donor 013-D who had a 12-fold increase (from 1/90 to 1/1122). Overall, 24/36 (67%) and 12/36 (33%) donors had either sustained or declined neutralizing antibody titers, respectively, during the observation period compared with their individual baseline titers.

**Figure 4.**
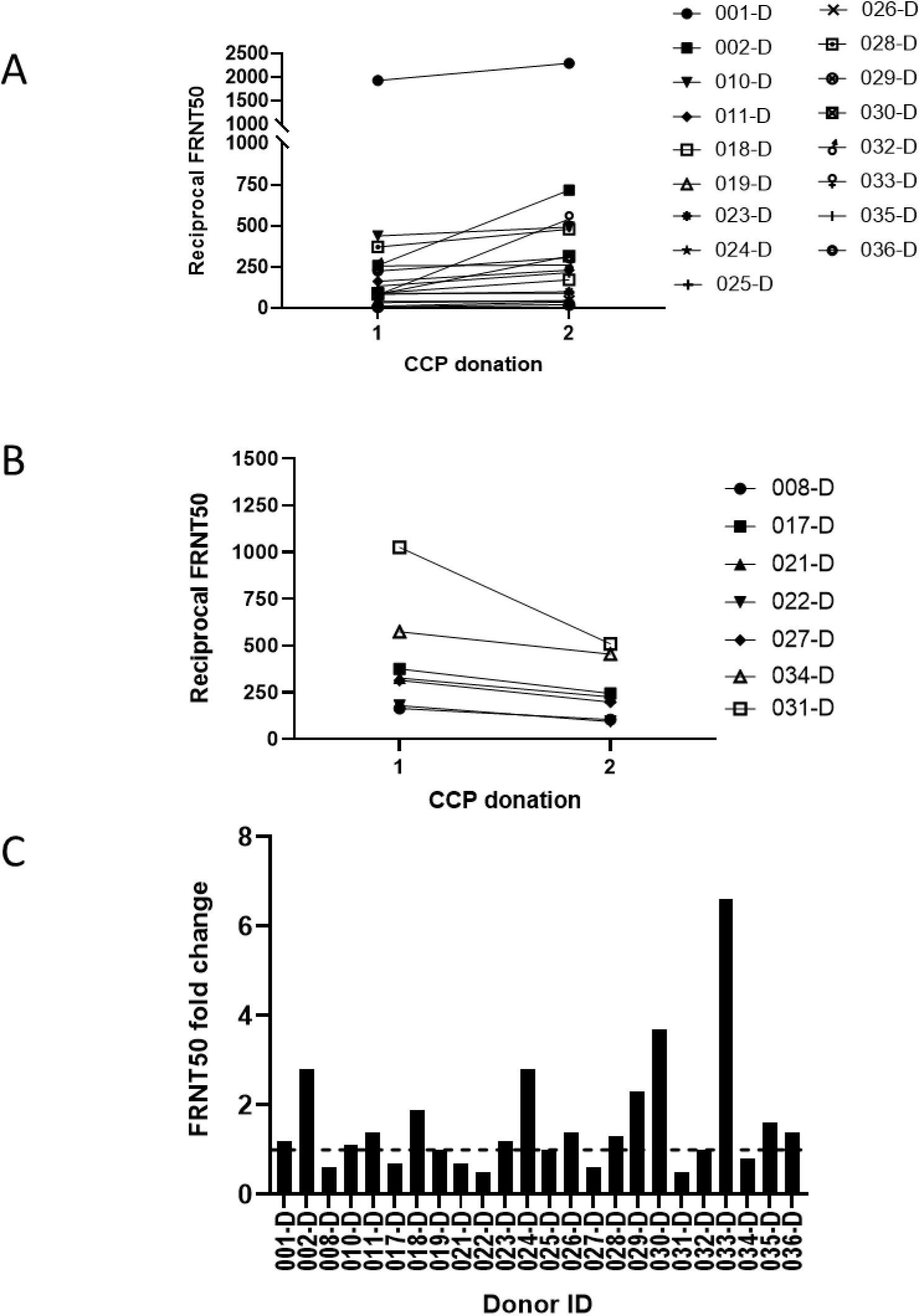
Change in FRNT50 titers over two sequential donations. FRNT50 titer increased or remained relatively unchanged (A, n=17 donors). FRNT50 titers decreased from the initial to the second donation (B, n=7). Fold increase from initial donation for donors with two sequential donations (C, n=24).

**Figure 5.**
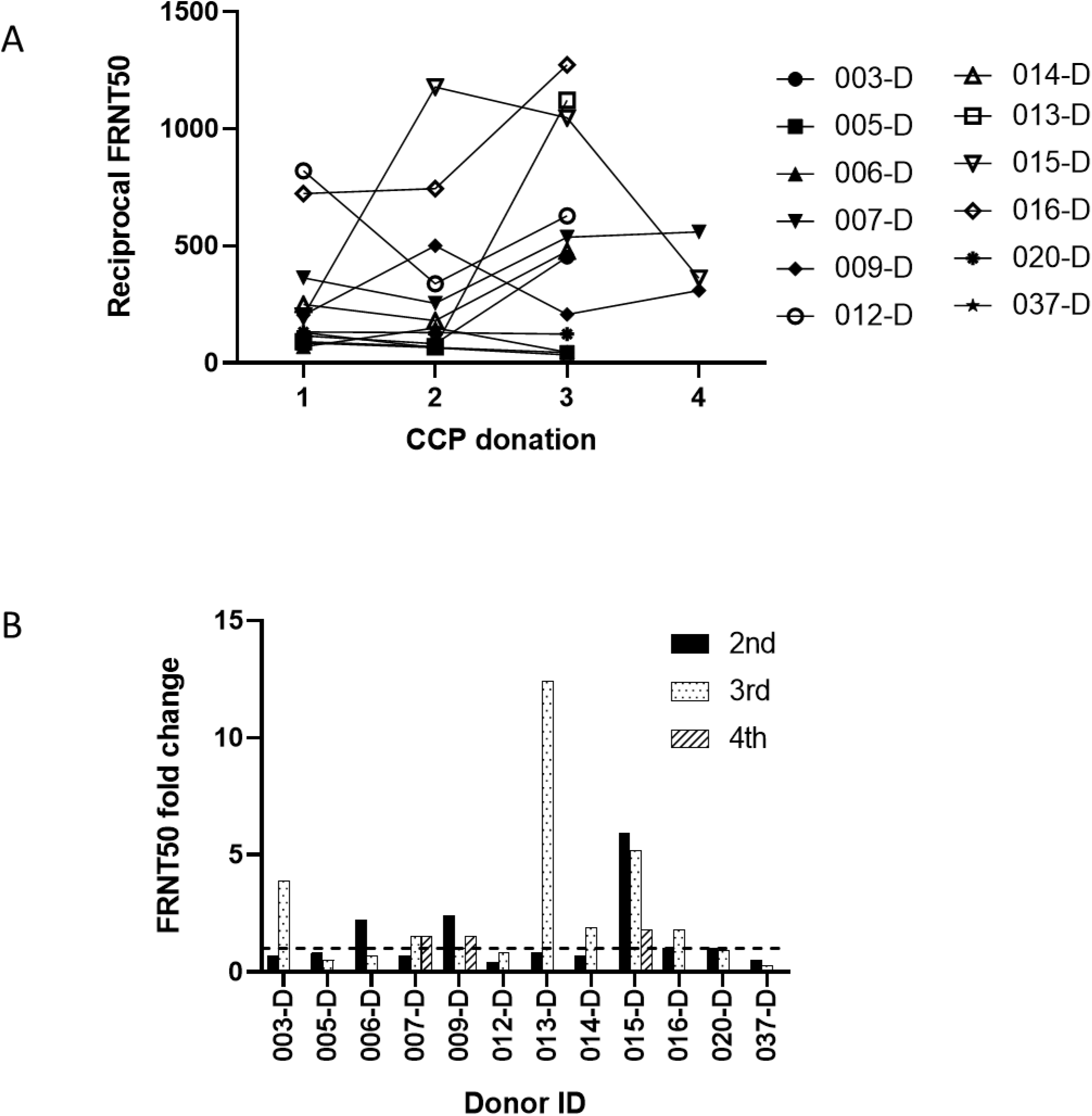
Change in FRNT50 titer over three or four sequential donations. (A) FRNT titers over the course of three or four donations for individual donors. (B) Fold-increase from initial donation. n=12.

## Discussion

Passive transfer of convalescent plasma has been utilized to combat infection with a variety of pathogens including the 1918 H1N1 influenza virus (9), the 2009 H1N1 influenza virus (10), and the SARS-CoV-2 -related coronaviruses, MERS and SARS-CoV (11, 12). Transfer of CCP for the treatment of COVID-19 received FDA approval for use as an investigational therapeutic in March of 2020 (13). Data regarding its efficacy continues to accumulate, and some initial successes have been reported (14-19). We have collected 548 units at CHCO to date. As this program ramps up to meet the growing need for CCP to combat the COVID-19 crisis, there is a need to screen these units as quickly and as accurately as possible for titer and quality of SARS-CoV-2 antibodies.

A large variety of serological assays for analysis of SARS-CoV-2 antibodies targeting the immunodominant nucleocapsid and spike antigens are now available with varying levels of regulatory approval and validation. However, these antigen binding assays do not provide information on neutralizing capacity of SARS-CoV-2 antibodies. Given that the FDA’s recommendation for CCP is a neutralizing antibody titer of at least 1:80 and preferably 1:160, and that neutralizing antibody assays cannot be easily implemented in most clinical laboratories, it is important to evaluate correlations between antigen binding assays and neutralizing antibody titers in order to triage CCP units and enable selection of units that are likely to have recommended neutralizing titers. An analysis of 159 serum samples from healthy, COVID-19 recovered individuals revealed that while samples with high IgG ELISA titers to the RBD antigen generally correlated with neutralizing activity as determined by a PRNT assay, only half of the individuals tested had >1:160 PRNT50 titer (20). Our comparison of anti-N IgG antibodies, measured by the EDI assay, and anti-S1-RBD IgG antibodies, measured by the Euroimmun assay, indicates that an increase in the relative level of positivity of either antibody correlated with an increase in neutralization titer, however, these correlations were mutually exclusive. The level of positivity for S1-RBD antibodies had a higher correlation with neutralizing activity, as would be expected, since viral entry is mediated by the spike protein and anti-N antibodies are non-neutralizing (21, 22). SARS-CoV-2 N antigen is highly antigenic (23), and while some of the donor samples were highly positive for anti-N IgG antibodies, they did not have corresponding high neutralizing antibody titers. The drawback of these qualitative assays is that they are based on a single dilution and, therefore, may not measure antibodies in the linear range. Despite this, our data indicate that samples with an anti-N IgG OD_450_ of 0.4 by the EDI assay, or a ratio of 3.0 for measurement of anti-S1-RBD IgG antibodies by the Euroimmun assay accurately captured the majority of samples with neutralizing titers of ≥1:80. Furthermore, our data suggest that analysis of antibodies to a single SARS-CoV-2 antigen may not be sufficient to be predictive of neutralizing capability.

There is limited information about the peak and decline of SARS-CoV-2 antibodies following generation of an antibody response. In a scientific brief released by the World Health Organization (WHO), it was stated that there is ’’currently no evidence that people who have recovered from COVID-19 and have antibodies are protected from a second infection” and further stated that laboratory tests that detect antibodies to SARS- CoV-2, including rapid immunodiagnostic tests, need further validation to determine accuracy and reliability (24). The presence of antibodies and how long they last in an individual may be a critical factor as the economy continues to reopen and people who are considered immune may be deemed safe to return to work or other activities (WHO). In this context, antigen binding assays that have been implemented for rapid screening do not provide information regarding the functionality of these antibodies. Such information is typically generated from biological assays such as the FRNT50, described in our study, that examines the ability of CCP to neutralize viral replication in permissive cell lines. We found that close to 80% of the population of donor samples we tested had a neutralizing antibody titer of ≥1:80, and 60% ≥1:160, both of which meet the FDA’s criteria for eligible CCP units.

The longevity of the antibody response is a critical part of potential protection against reinfection, although such information continues to be gathered. Analysis of the longevity of the antibody response to SARS1 indicates that anti-SARS1 antibodies were detectable two to three years following infection in one study (25), and, in a second study, detectable for close to a year following infection but declined over the course of this observation period (26). Short term studies on the durability of the SARS-CoV-2 antibody response have provided variable results, and in particular seem to correlate with the severity of illness (27) while other reports suggest antibody titers may quickly wane, and concerns for reinfection, particularly with mild or no symptoms, are not unwarranted (28). For example, one reported case found that a SARS-CoV-2 confirmed infected patient’s IgG antibodies became undetectable by day 80 (29). The significance of a waning antibody level may be particularly impactful when considering repeat collections from such donors, because of the effect that a waning antibody response may have on the amount and quality of the SARS-CoV-2 antibodies that are transfused as part of CCP therapy. As reported in the large study of 35,322 patients who received convalescent plasma as part of the Mayo Expanded Access Protocol, a higher antibody titer was correlated with reduced mortality (19). As a result, the recently issued Emergency Use Authorization from the FDA has established a cut-off value for a high antibody titer on the FDA approved Ortho VITROS SARS-CoV-2 IgG platform (30). Although the Signal-to-Cutoff ratio was provided for these reports, a correlation to neutralizing antibody titer thresholds in the prior FDA requirement of >1:80 titer with preference for ≥1:160 titer was not provided. This incites the question of the adequacy of the minimum threshold for CCP treatment and may impact future collections if the threshold is increased for therapeutic efficacy.

Our data suggests that a majority of donors (67%) had a neutralizing antibody response that was either sustained or increased over the short period of approximately three weeks to two months following a positive SARS-CoV-2 PCR result, and a smaller percentage (33%) showed a decrease in neutralizing antibody titer over sequential donations. Notably, repeat donations did not appear to affect antibody titer for the majority (67%) of donors. A drawback of our dataset is that for the majority of donors, we were able to test only two time points (7-24 days apart) following a positive SARS-CoV-2 PCR test, making it challenging to comment on longer term sustainability of the response. The longevity of the SARS-CoV-2 antibody response and the level of protection it will provide for reinfection is yet to be determined.

## Data Availability

All data included in this manuscript was generated at Children's Hospital, Colorado and the University of Colorado School of Medicine

## Authorship Contributions

KA and MD developed the CCP donor program and contributed to writing and critical review of the manuscript. TEM and MKM developed the FRNT50 assays and contributed to writing and data analysis, MKM performed the FRNT50 assays and FRNT50 data analysis, LH, PAM and GA performed the EDI and Euroimmun ELISAs, analyzed ELISA data and contributed to writing, VK conceptualized the manuscript, contributed to writing, data analysis and critical review of the manuscript.

